# Effect of Umbilical Cord Milking on Severity of Hypoxic Ischemic Encephalopathy in Asphyxiated Neonates – A Pilot study

**DOI:** 10.1101/2021.05.26.21257569

**Authors:** Roshith. J. Kumar, V.C. Manoj

**Author notes:** **Correspondence to:** Dr V C Manoj, Professor and Head, Department of Neonatology, Jubilee Mission Medical College, Thrissur, Kerala – PIN – 680005, E mail.

## Abstract

**Background:** The present study was aimed to evaluate the effect of umbilical cord milking technique on severity of Hypoxic Ischemic Encephalopathy (HIE) in asphyxiated neonates and assessed by Modified Sarnat’s staging as primary outcome, APGAR score at 5 minutes and Respiratory support requirement as secondary outcome.

**Methods:** This was a randomized, controlled pilot study conducted in neonatology department at a tertiary care centre, Thrissur, Kerala for one year starting from March 2020. The neonates were divided into two groups non milking group, control (n=38) and umbilical cord milking, case [UCM] (n=32) and their outcomes were compared. In the intervention group, the cord was cut at 30 cm from umbilical stump within 30 seconds of birth and euthermia was maintained. The umbilical cord was raised and milked from the cut end towards the infant 3 times with speed at 10 cm/sec and then clamped 2-3 cm from the umbilical stump. In the control group, the umbilical cord was clamped without doing cord milking.

**Results:** In this study moderate to severe HIE were less in case group 46.9 % than control group 55.1% and less neonates 44.7 % had Mild HIE in control group compared to case group 53.1% even though result was statistically not significant as primary outcome (p value – not significant). Eight neonates (21.6%) in control group had Apgar at 5 min score 0-3, whereas only 4 (12.5%) neonates in cord milking group.

**Conclusions:** The insufficient knowledge of placental transfusion limits and benefits leads to a wide variation in the management of cord clamping. It would be useful to standardise the UCM procedure in order to offer protocols applicable to clinical practice, and to spread knowledge among professionals through educational programs.

## INTRODUCTION

Neonatal Resuscitation Protocols (NRP 2015 & 2020) recommend delayed cord clamping (DCC) as part of normal neonatal resuscitation, after International Liaison Committee on Resuscitation (ILCOR) systematic review found that interventions to enhance placental transfusion like DCC are beneficial in neonates. DCC is associated with less intraventricular haemorrhage, higher blood pressure and blood volume, less need for transfusion after birth, and less necrotizing enterocolitis.^1^ Umbilical Cord Milking (UCM), another intervention to enhance placental transfusion has been shown to be a safe procedure that improves the haemoglobin and iron status at 6 weeks of life among term and late preterm neonates.^2,3^ In a comparison of two types of intervention (DCC and UCM) to enhance placental transfusion in term infants, there was no difference in haemodynamic status, Cranial Doppler indices, and adverse neonatal outcomes among the two groups.^4^ Currently Neonatal Resuscitation Protocols (NRP 2020) states that cord milking is being studied as an alternative to delayed cord clamping but should be avoided in babies less than 28 weeks gestational age, because it is associated with brain injury.^5^

Birth asphyxia accounts for 20-30% of neonatal deaths in India and surviving infants are at risk for development of cerebral palsy and neuro developmental disabilities in later life. Hypoxic Ischaemic Encephalopathy (HIE) is a brain injury seen in these neonates due to inadequate blood flow and oxygen delivery to the brain. Currently only therapeutic hypothermia (whole body temperature maintained at 33.5°C) initiated within 6 hours of birth and continued for 72 hours is known to be effective in preventing secondary neuronal injury in neonates with moderate to severe HIE. UCM may act as an adjunctive to cooling and can potentially improve brain injury in neonates with HIE.^6,3^

UCM is proposed as a simple, safe intervention in depressed neonates^8^ which if found to be beneficial, could be recommended as a standard protocol for all depressed neonates in resource limited settings like primary and community health centre. UCM could then reduce the need for neonatal resuscitation interventions (positive pressure ventilation, chest compressions, medication and fluid boluses). It may also prove to be a useful adjunct to whole body hypothermia for those neonates with moderate to severe HIE. This simple intervention can be widely adopted in both developed and developing countries, thereby helping the latter to achieve the Millennium Development Goals.

Placenta being a rich source of stem cells, UCM may potentially improve brain injury in neonates with HIE. UCM may prove to be a relatively harmless and simple intervention feasible among the initial steps in the resuscitation of a depressed newborn.^9^ If found to be beneficial in preventing brain injury, it has immense therapeutic value in resource limited settings like primary and community health centre, where birth asphyxia otherwise carries a very poor outcome.

The present study was conducted to investigate severity of HIE of Umbilical cord milking in term infants who are depressed at birth using Modified Sarnat Score.

Later investigations such as APGAR score of 0-3, 4-6 & 7-10 at 5 minutes, respiratory off support at 48 hours, Non Invasive Mechnanical Ventiltation (NIMV) & Invasive Mechanical Ventiltation (IMV) requirement and MRI abnormality were evaluated on the study participants.

## MATERIALS AND METHOD

This was a randomized, controlled pilot study conducted in neonatology department at a tertiary care centre, Thrissur, Kerala for one year starting from march 2020. The study was approved from the Institutional Ethical Committee. The trial was registered under the ClinicalTrials.gov (ClinicalTrials.gov Identifier: NCT03123081). All depressed neonates (defined as per NRP 2015 & 2020 criteria) of gestation 35 weeks and above, delivered either vaginally or by lower segment caesarean section with in the hospital were enrolled into the study after receiving informed consent from the parents. Cases such as MCDA Twin pregnancy (including DCDA twins), triplet or quadruplet pregnancy, short umbilical cord length (< 25 cm), hydrops foetalis, major chromosomal or congenital anomalies, severe placental abruption and cord prolapse and cord abnormalities such as true knots were excluded from the study.

### Randomization

A computer generated random number of all term depressed neonates were allocated to intervention and control group. Consent of all term parents were taken before delivery.

Ethical clearance no:01/17/IEC/JMMC&RI

### Details of the intervention

All doctors and nurses involved in delivery and newborn resuscitation were trained for a standardized cord-milking technique, by showing three live demonstrations by the principal investigator and also showing a video available on the internet, by Tarnow Mordi, *et al*.^10^ In all cases after birth, the babies were held at the level of the uterus in vaginal delivery and on the thighs of mother in caesarean section while the umbilical cord was cut and clamped.

In the intervention group, the cord was cut at approximately 30 cm of length from umbilical stump within 30 seconds of birth (early clamping). Then the baby was placed under the radiant warmer for resuscitation as per NRP 2015 guidelines. The umbilical cord was raised and milked from the cut end towards the infant 3 times with speed at 10 cm/sec and then clamped 2-3 cm from the umbilical stump.

In the control group, the umbilical cord was clamped early (within 30 seconds) near the umbilicus and cut without doing cord milking.

The primary outcome of the study was to investigate the severity of HIE of UCM in case and control group using Modified Sarnat Score. Secondary outcomes were to investigate APGAR score of 0-3, 4-6 & 7-10 at 5 minutes, respiratory off support at 48 hours, Non Invasive Mechnanical Ventiltation (NIMV) & Invasive Mechanical Ventiltation (IMV) requirement and MRI abnormality on the study participants.

### Statistical Analysis

Data were analyzed using Chi-square test, student t – test, percentage anal6ysis, odds ratio. Means were compared using the t-test. Adjustments for multiple comparisons were made using Fisher’s least significant difference method. Adjusted were obtained using analysis of covariance, and differences were compared using Fisher’s least significant difference t test.

### Flow Diagram

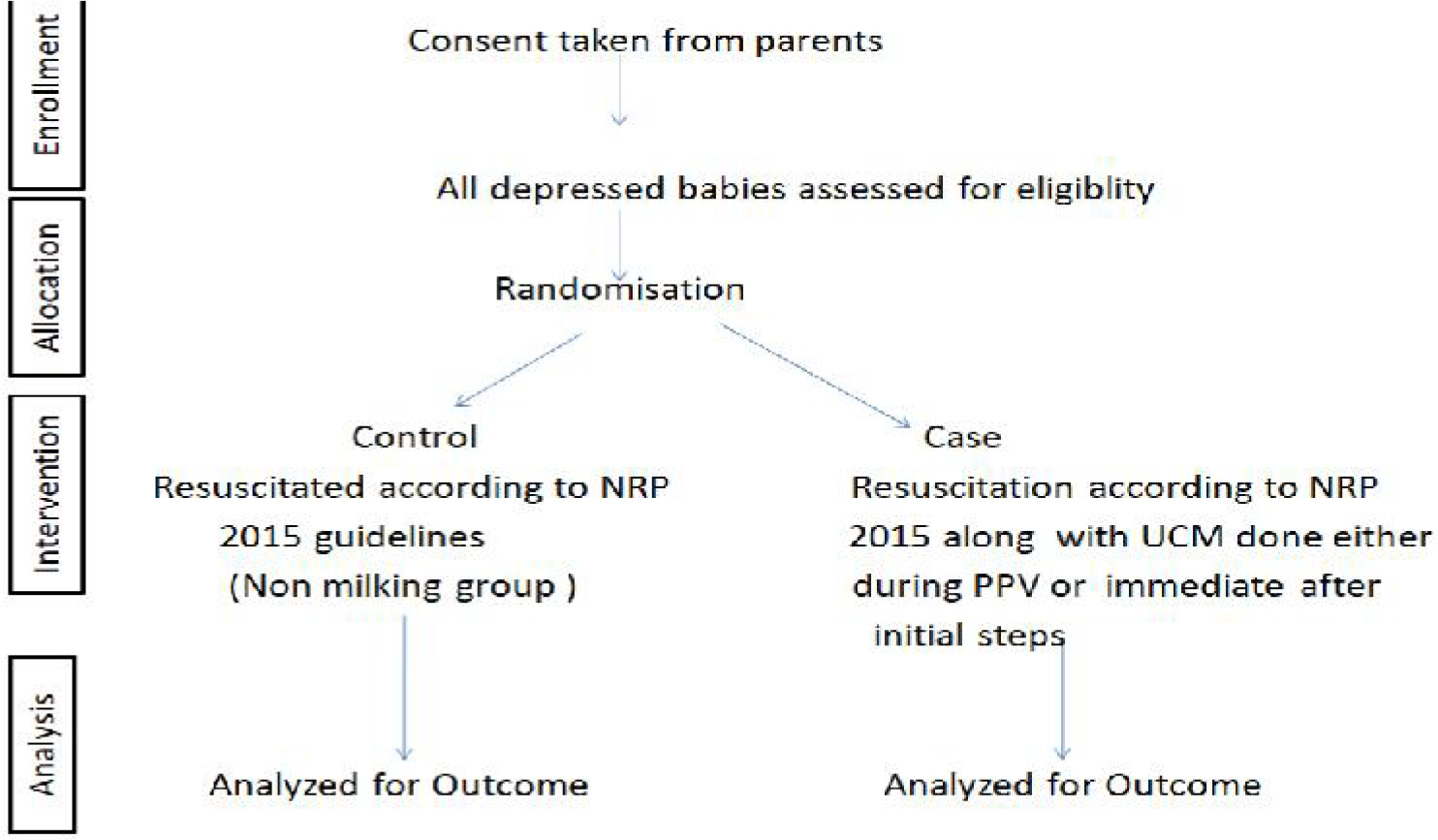

## RESULTS

A total of 70 babies were recruited for this study that fulfilled the inclusion criteria. These babies were randomized in to two groups: intervention group (38) and control group (32). The baseline characteristics of the two groups were compared in 70 new born enrolled for the study (Table 1).

**Table 1:**
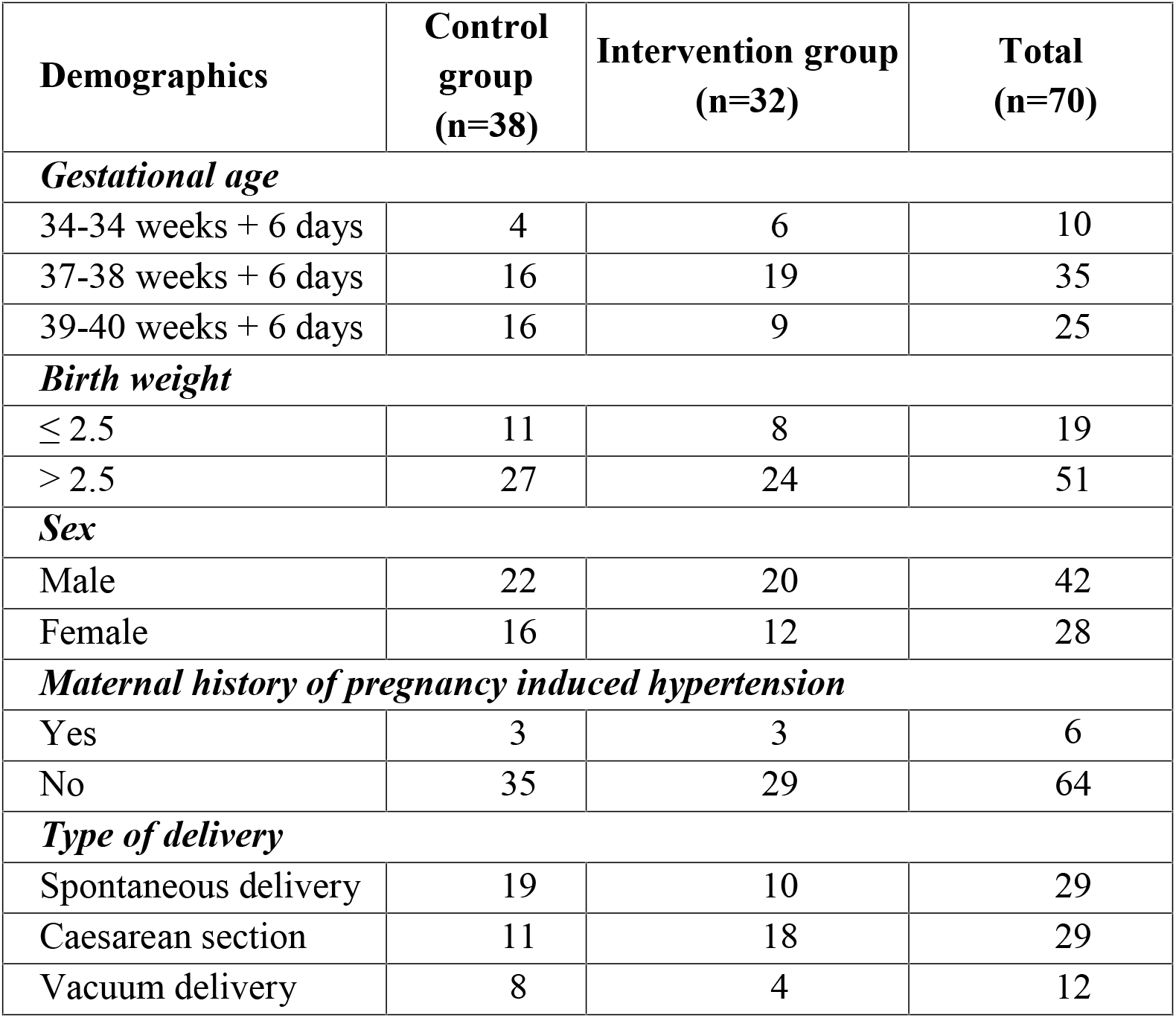
Baseline characteristics.

When primary outcome was compared in both the groups, it was observed that moderate HIE was present in 46.9 % in intervention group as compared to 55.3 % in control group, indicating a trend towards better outcome in intervention group (p value – not significant).

When secondary outcomes were compared (Table 2), in intervention group, 12.5% had APGAR score of 0-3, 53.1% had score of 4-6 and 34.4% had score of 7-10 where as in control group more neonates had score of 0-3 21.6 %, 43.2 % had score of 4-6 and 35.1% neonates had scoreof 7-10. Among the intervention group, 28.1% required off support at 48 hours and in control group 13.2% required off support at 48 hours. In the present study, 52.6% required noninvasive ventilation, 39.5 % required mechanical ventilation and 7.9% did not required respiratory support in control group, whereas in Intervention group 56.3 % required noninvasive ventilation and 40.6% neonates required mechanical ventilation. Among the intervention group, 26.7% showed abnormal MRI and in control group 64.7% showed abnormal MRI.

**Table 2.** Primary & Secondary outcomes.

| Table 2.Primary & Secondary outcomes |  |  |  |  |  |  |
| --- | --- | --- | --- | --- | --- | --- |
| Variables | Category | Milking of cord |  |  |  | P Value |
|  |  | Not done (control) |  | Done (case) |  |  |
|  |  | n=38 | Percentage | n=32 | Percentage |  |
| Primary outcomes |  |  |  |  |  |  |
| HIE | No HIE / mild HIE | 17 | 44.70% | 17 | 53.10% | 0.484 |
|  | Moderate to Severe HIE | 21 | 55.30% | 15 | 46.90% |  |
| Secondary outcomes |  |  |  |  |  |  |
| APGAR at 5' | 0--3 | 8 | 21.60% | 4 | 12.50% | 0.556 |
|  | 4--6 | 16 | 43.20% | 17 | 53.10% |  |
|  | 7--10 | 13 | 35.10% | 11 | 34.40% |  |
| Off support at 48hrs | No | 33 | 86.80% | 23 | 71.90% | 0.119 |
|  | Yes | 5 | 13.20% | 9 | 28.10% |  |
| NIMV/IMV | Off support | 3 | 7.90% | 1 | 3.10% | 0.691 |
|  | NIMV | 20 | 52.60% | 18 | 56.30% |  |
|  | IMV | 15 | 39.50% | 13 | 40.60% |  |
| MRI | Normal | 6 | 35.30% | 11 | 73.30% | 0.031 |
|  | Abnormal | 11 | 64.70% | 4 | 26.70% |  |

## DISCUSSION

There are no recent clinical evident trails to emphasize the importance of placental transfusion in neonatal outcomes and the role of each component of placental transfusion believed in the past by the WHO for the active management of the third stage of labour concepts were changed. Hence, the practice of immediate cord clamping was excluded by the WHO guidelines in 2012, and cord traction was defined as optional.^12,13,14,15^ Placental transfusion is the transfer of placental blood to neonate during the first few minutes of life.^16^ This procedure helps in reducing rates of mortality in asphyxiated neonates and prevent iron deficiency anaemia in term neonates.^17,18^

Placental transfusion conducts through DCC or UCM, may also acts as an important procedure for ensuring better neonate outcome without a stormy course of NICU stay. DCC and UCM both helps in increasing arterial oxygen content, maintain haemodynamic stability and can be also done in low-resource settings. Cord clamping time, uterine contractions, umbilical blood flow, breathing, and gravity all play a central role in determining placental transfusion effectiveness.^16^

DCC provides a passive transfer of placental blood, at a slow rate meanwhile UCM is an active stripping of blood through the umbilical cord to the neonate, as a faster method.^3^

According to RCOG article, 2015, UCM is an alternative to DCC in case of preterm births, but it needs to be further investigated in order to evaluate associated benefits and risks so that it can be performed world wide.^19^

There is a need for larger randomised trials on UCM to be conducted in developing countries and has insufficient knowledge to prove its neonatal outcome. Moreover, there is no standardisation on DCC method and its optimal time to do, despite of time is being essential for passive placental transfusion. As it is common practice is to give a depressed newborn to the neonatologist as soon as possible after birth considering the safety of depressed neonates who requires resuscitation in first golden minute.

According to Italian recommendations in case of ceaseran term newborns, if DCC cannot be performed, UCM may be considered as an alternative procedure with the purpose of increasing haemoglobin levels in postnatal period and iron reserves in the following weeks.^22^

In fact, UCM is believed to be a simple procedure that can be safely performed in a matter of seconds also by obstetricians, with no time. Furthermore, this method may also be very useful in cases of neonatal asphyxia, helping in the crucial importance of neonatal outcomes. Girish et al, UCM may be a feasible procedure can be done in depressed neonates along with resuscitation, it will not cause any resuscitation delays or harm in depressed newborns compared to ICC.^10, 21^

Katheria et al. also concluded in their retrospective analysis that neonates with acidosis, who had received UCM and needed resuscitation and ongoing respiratory support were fewer in number than those who received ICC.^23^ There are ongoing cited studies evaluating haematological parameters such as haemoglobin, haematocrit, and ferritin which can prove that there is significantly higher UCM results compared to ICC groups at different times from the delivery.^18,22,23,24^ Inspite of having good number of studies evaluating UCM outcomes, a standardised procedure for milking is needed and it should be followed internationally. McAdams’s is the only study evaluating different outcomes between different UCM procedures, showing that I-UCM (×3 or ×4) promotes a larger transfusion of blood volume to newborns at birth than C-UCM.^3^ Even if published data on UCM enlightens positive effects of encouraging, stating that UCM may be the most effective method to provide placental transfusions and UCM received depressed neonates would have lesser NICU stormy course.

The implementation of procedure could also be associated with clinically approved UCM guidelines availability, knowledge of UCM in short and long term neonatal outcomes, and requires friendly cooperation within the delivery team.

In conclusion, there is insufficient knowledge and inappropriate practice to conduct placental transfusion within in the limits and its benefits leads to a world wide variation in the managing of cord clamping. It would be useful if UCM practice is standardized in all depressed neonates at birth, included in third stage of labour management and its guidelines to spread knowledge among professionals through educational programs and conferences.

### Limitations

Since this being a pilot study smaller sample size, co-morbidities could not be separated, long term outcome not assessed, chance error because of lack of blinded randomisation could affected outcome.

## Supporting information

Ethics certificate & list of members

## Data Availability

Not applicableNo funding

## Abbreviations

HIE: Hypoxic Ischemic Encephalopathy
UCM: Umbilical Cord Milking
NIMV: Non Invasive Mechanical Ventilation
IMV: Invasive Mechanical Ventilation

## Acknowledgement

The authors thank Ms. Ditty Martin, Research Assistant, Neonatology Department, JMMC & RI for providing help to filter data from hospital records and to Ms Mridula Vellore, Research Assistant (Scientific writing) for editing the article.

