## Supplementary material for "Effect of Umbilical Cord Milking on Severity of Hypoxic Ischemic Encephalopathy in Asphyxiated Neonates – A Pilot study": Ethics certificate & list of members

### JUBILEE MISSION MEDICAL COLLEGE & RESEARCH INSTITUTE

THRISSUR 680 005, KERALA

### Institutional Ethics Committee Certificate

*The Institutional Ethics Committee, Jubilee Mission Medical College, Thrissur has gone through the details of the Clinical research work*

Ref. No : 01/17/IEC/JMMC & RI
Name : DR. VARANATTU C MANOJ
Course : MD/MS/DNB/Other (Specify)
Department : NEONATOLOGY
Title : ROLE OF UMBILICAL CORD MILKING IN THE MANAGEMENT OF HYPOXIC-ISCHEMIC ENCEPHALOPATHY IN NEONATES: A RANDOMIZED CONTROLLED TRIAL
Name of ~~Guide~~/ Co- ~~Guide~~ : INVESTIGATORS: DR. BINDU MENON/ DR. ASHWATH KUMAR
Approval Date : 04.01.2017

*and has decided to give permission to proceed with the study / publish the article.*

*Researchers shall inform the Institutional Ethics Committee,*

On completion of the study

If any difficulty may arise during the clinical trial or

If the study is dropped quoting the reason.

*Secretary*
SECRETARY
ETHICS COMMITTEE
JMMC & RI, THRISSUR 5
*Date :*..........

*Chairperson*
CHAIRPERSON
ETHICS COMMITTEE
JMMC & RI, THRISSUR
*Place :*..........

**File No. ECR/1411/Jubilee/Inst/KL/2016**

**Government of India**
Ministry of Health & Family Welfare
Directorate General of Health Services
Office of Drugs Controller General (India)
Central Drugs Standard Control Organization

**FDA Bhawan, Kotla Road,**
**New Delhi – 110 002, India**
**Dated:** 05/08/2016

**To,**

**The Chairman**
**Institutional Ethics Committee,**
**Jubilee Mission Medical College & Research Institute,**
**P. B. No. 737, Bishop Alappatt Road, Jubilee Mission P. O.,**
**Thrissur East-680005, Kerala, India.**

**Subject:** Ethics Committee Registration No. **ECR/835/KL/Inst/2016** issued under Rule 122DD of the Drugs & Cosmetics Rules1945

**Sir/Madam,**

Please refer to your application no. JMMCIECDCGI/2014/001 dated 12.12.2015 submitted to this office for the registration of Ethics Committee.

Your Ethics Committee is hereby registered under Rule 122DD vide Registration No. **ECR/835/KL/Inst/2016** with the following composition and all the condition mentioned under the Registration certificate issued to you.

| Sr. No. | Name of member | Qualification | Role/Designation in Ethics Committee |
|---|---|---|---|
| 1. | Dr. Zacharias Cherian | M.Sc (Pharmacology), B.V.Sc | Chairman |
| 2. | Dr. M.P. Raphael | MD (Pharmacology) | Member Secretary |
| 3. | Dr. K.B. Mohan | MD (General Medicine) | Clinician |
| 4. | Dr. Annie Kuriyan Thadicaren | MD, DGO | Clinician |
| 5. | Dr. C.V. Andrews | MS (Ophthalmology) | Clinician |
| 6. | Dr. Mariam Koshi Thomas | MD, DA (Anesthesiology) | Clinician |
| 7. | Dr. V.K. Ramankutty | MD (Forensic Medicine) | Clinician |
| 8. | Dr. Alex Joseph | MS (Ophthalmology) | Clinician |
| 9. | Dr. M.V. Suresh | MS (General Surgery) | Clinician |
| 10. | Dr. Biju Bahuleyan | MD (Physiology) | Basic Medical Scientist |
| 11. | Adv. Anto Davis A | B. Sc, LLB | Legal Expert |
| 12. | Mr. Sijo Jose | MSW | Lay Person |
| 13. | Fr. Baiju Chakkery | BA, MSW | Social Scientist |
| 14. | Dr. Kumudam Unni | B.Sc, PhD | Scientific Member |
| 15. | Dr. Angela Gnanadurai | M.Sc, PhD | Scientific Member |

**(Dr. G. N. Singh)**
**Drugs Controller General (I) & Licensing Authority**

Dr. G. N. SINGH
Drugs Controller General (India)
Dte. General of Health Services
Ministry of Health & Family Welfare
FDA Bhawan, Kotla Rc [illegible] I.T.O.
New Delhi-110002
